# Personality factors and childhood adversity in psychiatric patients with and without recent suicide attempts: a cross-sectional study

**DOI:** 10.64898/2026.05.25.26354029

**Authors:** Lejla Colic, Johanna Mußlick, Ani Zerekidze, Lydia Bahlmann, Benjamin Buske, Martin Walter, Fabrice Jollant, Gerd Wagner

## Abstract

**Background:** Childhood adversity (CA) is recognized as a distal risk-factor for suicide attempts (SA) in individuals with psychiatric disorders. However, not all individuals with experiences of CA will engage in SA. Contributing to this relationship may be proximal factors such as impulsivity, inward anger and self-aggression. However, these factors are often conceptually blended and measured in different samples. We sought to clarify association among CA and personality factors in persons with SA.

**Methods:** Participants from two studies comprised individuals with a diagnosed psychiatric disorder and history of SA (n= 139) and individuals with depressive disorder (clinical controls, CC; n= 24). We investigated self-reported levels of CA, impulsivity, inward anger, and self-aggression between the SA and CC (p_corr_< .012). We tested the relationship among the factors using regression (p_corr_<.017) and mediation model (indirect effects, p<.05) within the SA group. Sensitivity models were run controlling for age, gender, symptom severity, trait anger, and externally oriented aggression.

**Results:** SA group had higher impulsivity (p_corr_=.067) in a model controlled for age and gender. Other factors did not differ among groups. Within the SA group the analyses revealed positive association among CA and personality factors (p_corr_’s<.06) in basic and model with age and gender, however the association was not specific for internally (self) oriented factors (coefficient comparison, p<.07). Parallel mediation model indicated that CA had indirect effect on self-aggression through impulsivity (p=.001) and to a lesser extent through inward anger (p=.066). Generally, models controlling for cognitive depression symptoms showed less prominent effects (p_corr_>.1).

**Limitations:** The study was cross-sectional and did not include behavioral tasks (state) measures of proximal factors.

**Conclusions:** CA and personality factors showed similar severity levels among the SA and CC groups suggesting they may relate to broader psychopathologies, rather than specifically to SA. The association of CA with anger and aggression was unspecific to internally oriented factors indicating the need for more precise measuring instruments developed specifically for individuals with SA. Overall, the study highlights personality factors as being associated with risk in broader vulnerable populations.

## Introduction

The World Health Organization estimates that around 700 000 people annually die by suicide (World Health 2025). A recent review highlighted the observation that suicidal behavior (SB) occur across psychiatric diagnoses (Xu et al. 2023). Supporting this notion, several theoretical models have been proposed to explain the transition to SB, regardless of psychiatric disorder, emphasizing the roles of both distal and proximal risk factors (Van Orden et al. 2010, Baumeister 1990, O’Connor 2011, Klonsky and May 2014, Mann and Rizk 2020).

There is evidence that childhood adversity (CA) is among the strongest distal risk factors for suicidal behavior (SB) (Turecki 2014, Alfonso and Schulze 2021). Meta-analyses have shown that CA, whether measured retrospectively (Angelakis, Austin, and Gooding 2020) or prospectively (Zatti et al. 2017), is associated with a higher risk for SB across populations. Although many individuals who experience CA, such as physical, sexual, or emotional abuse, do not go on to exhibit SB, these forms of adversity are strongly linked to suicide attempts (SA) and death by suicide (Baldini et al. 2025, Castellví et al. 2017).

Exposure to CA disrupts normative emotional development, often leading individuals to develop maladaptive strategies such as internalizing rather than processing and expressing negative affect (Harms, Leitzke, and Pollak 2019, Milojevich, Lindquist, and Sheridan 2021). Within the framework of the *Three-Step Theory* (Klonsky and May 2015) CA is acts as a distal risk factor, while the maladaptive strategies are proximal factors that contribute to the emergence of the suicide ideation (SI) (Laghaei et al. 2023). These maladaptive may include inward-directed anger, which can manifest as self-aggressive cognitive patterns, such as intense self-criticism, shame (Jang et al. 2014, Lin et al. 2022) or impulsive behavioral patterns aimed at controlling internal states (Cyders and Smith 2008). Over time, these maladaptive strategies can increase psychological distress, with self-aggression serving as both a marker of distress and a potential mechanism that habituates individuals to self-harm (O’Donnell, House, and Waterman 2015), thereby lowering the threshold for transition to SB (Turecki et al. 2019).

These maladaptive strategies, namely impulsivity related to affective states, inward anger, and self-aggression are multidimensional and vary in expressions, making it challenging to clearly delineate and assess their contributions to SB thereby increasing the risk of conceptual and measurement overlapping.

Impulsivity ranges from spontaneous risk-taking to difficulties resisting urges, each associated with distinct neurobiological pathways (e.g., (Dalley and Robbins 2017)). It can be assessed using complementary self-report measures and laboratory-based cognitive tasks (Dalley and Robbins 2017, Sharma, Markon, and Clark 2014). To clarify and assess trait impulsivity as a multidimensional construct, Whiteside and Lynam (2001) developed the Impulsive Behavior Scale based on the Five-Factor Model of personality. Using an exploratory factor analysis of impulsivity self-report measures, they derived a four-factor model of impulsivity (i.e., UPPS) (Whiteside and Lynam 2001).

Childhood adversity has been associated with various facets of impulsivity (Liu 2019, Aubrey, Jones, and Paddock 2025) suggesting that impulsivity may mediate relationship between CA and SA. In SA research, urgency, a scale to measure impulsivity is a particularly interesting impulsivity measure as it may be relevant for general negative affectivity (Weitzman, McHugh, and Otto 2011, Bahlmann et al. 2025) (although for momentary associations see (King et al. 2024)). A recent meta-analysis found that that among self-reported impulsivity facets, negative urgency has a robust association with suicide-related outcomes (Bruno et al. 2023). However, the authors did not observe clear differences in UPPS dimensions between groups with SI and SA, pointing to the potential importance of additional moderating and/or mediating variables, such as personality-related traits, including inwardly directed anger and self-aggression, in the transition from SI to SA.

Similar to impulsivity, anger and aggression have been conceptualized in various ways, each linked to different biopsychosocial pathways (e.g. (Spielberger 2010, Allen, Anderson, and Bushman 2018)). Both anger and aggression have been individually associated with higher levels of CA (de Bles et al. 2023, Drachman et al. 2022, Simsek and Evrensel 2018) and have also been reported as risk factors for SA (Ammerman et al. 2015, Dillon, Van Voorhees, and Elbogen 2020, Wang et al. 2014, Lee et al. 2025).

Overall, both CA and specific personality factors have been reported to be elevated in individuals with a history of SA. However, the literature is marked by conceptual ambiguity and inconsistent definitions of impulsivity and the anger-aggression behavioral cluster. This issue is compounded by their high intercorrelations and overlapping operationalizations of these behavioral factors, which hinders the translation of observed associations to practical targets for psychotherapy. Specifically, studies have not differentiated cognitive impulsivity, marked by a lack of planning and reflection, from impulsivity rooted in negative affect. Similarly, distinctions remain blurred between trait and internally oriented anger, as well as reactive and self-aggression. In addition, studies have often compared SA groups to healthy or normative samples, which may conflate differences attributable to current psychopathologies.

To address these limitations, we investigated the associations between CA and specific personality factors, i.e., impulsivity, anger, and aggression, in a large sample of individuals with a well-defined history of *recent* (within the last year) SA. These factors were operationalized as urgency, inward anger, and self-aggression. The aims of our study were therefore threefold:

1) To investigate the differences between the SA group and clinical control group (CC), with a focus on the specificity of SA. We hypothesized that the SA group would show greater severity across all examined factors, particularly regarding CA severity and self-aggression severity. We additionally report the prevalence of CA subtypes measured with the short form of the Childhood Trauma Questionnaire (CTQ-SF) in the SA group in comparison to the German normative sample.
2) To examine, within the SA group, the association among CA, affective impulsivity, inward anger, and self-aggression, including sensitivity models controlling for depression severity, trait anger, and/or externally oriented aggression. We hypothesized a positive association between CA severity and these factors, with stronger relationships expected for self-oriented factors.
3) To test a theoretical mediation model within the SA group, with the CA as the predictor, affective impulsivity and inward anger as the mediators, and self-aggression as the outcome. We hypothesized positive regression paths among variables but made no specific hypotheses regarding the effect sizes.

## Methods

### Participants

The analysis sample was pooled across two studies: “Network for suicide prevention in Thuringia” (NeST) and “The choice of a violent suicidal means: a MRI study with computational modeling of decision-making” (SUICIDE_DECIDE). Both studies used the same inclusion criteria for the current suicidal behavior disorder (SBD) and same questionnaires.

In the NeST project adult inpatients with ages 18-60, all genders and fluent in German language, with a current SA were recruited from 2018 to 2020 in four different cooperating sites in Thuringia, Germany (Departments of Psychiatry and Psychotherapy of the University Hospital Jena, of the Thüringen-Kliniken “Georgius Agricola” Saalfeld, of the Asklepios Fachklinikum Stadtroda and the Helios Fachkliniken Hildburghausen). We only included participants who fulfilled the Diagnostic and Statistical Manual of Mental Disorders (DSM)-5 (APA 2013) criteria for the current SBD. In the DSM-5, “suicide attempt” is explicitly defined as “a self-initiated sequence of behaviors by an individual who, at the time of initiation, expected that the set of actions would lead to his or her own death”. Exclusion criteria were acute psychosis (based on DSM-IV), acute intoxication, withdrawal symptoms, diagnosed intelligence impairment, language barriers, lack of insight, and dementia diseases. The presence of psychiatric diseases was assessed by trained psychologists using the Mini International Neuropsychiatric Interview (M.I.N.I. 7.0) (Sheehan et al. 1998), a short structured diagnostic interview for DSM-IV Axis I disorders. NeST project was funded by the federal ministry of health (BMG, grant number: ZMVI1-2517FSB143). The local ethics committees of the Friedrich Schiller University Jena, and of the State Medical Association of Thuringia (a federal state in Germany) approved the study. All participants gave informed written consent.

The SUICIDE_DECIDE was done in the Department of Psychiatry and Psychotherapy, at the Jena University Hospital, Germany, and the Department of Psychiatry at the Nîmes Academic Hospital (CHU), France. The Friedrich Schiller University, Jena, Germany, and the *Comité de Protection des Personnes SUD-EST IV*, France approved this project. The study was registered on ClinicalTrials.gov (NCT05230043). The analysis in the present paper focuses on the sample collected at the Jena site. The data was collected between 2021 and 2022. Inclusion criteria for all participants were male or female gender, age between 25 to 60 years and fluency in German language. Inclusion criteria for the individuals with SA were fulfilling the DSM-5 criteria for current SBD. Inclusion criteria for the clinical control group were that they had a major depressive (MDD) or bipolar disorder (BD) but no personal or family (second degree) history of SBD. Only individuals that fulfilled the criteria for *current* depressive episode were included. The diagnoses for the groups were established using the M.I.N.I. 7.0, a short structured diagnostic interview for DSM-5 Axis I disorders (Sheehan et al. 1998). Exclusion criteria for all participants were current (hypo)manic or mixed episode and current psychotic disorder (based on DSM-5 criteria), withdrawal symptoms, acute intoxications, lack of insight, dementia, past brain trauma, and contraindications for magnetic resonance imaging. SUICIDE_DECIDE was funded by the American Foundation for Suicide Prevention (grant number LSRG-1-086-19).

### Psychometric assessment

Demographic data were collected with sociodemographic questionnaire that included questions on age, gender, basic socio-economic information, number of previous suicide attempts, family history of SB (in first-degree relatives), number of previous psychiatric or psychotherapy treatments, and current medication status.

<u>Childhood adversities</u> were measured with the German version of the short form of the Childhood Trauma Questionnaire (CTQ-SF; (Wingenfeld et al. 2010, Bernstein et al. 2003)). The CTQ-SF is a self-reported retrospective questionnaire that assesses physical, emotional and sexual abuse, and physical and emotional neglect. Each scale consists of five items rated with a five-point Likert scale (1-5). Overall good psychometric properties have been reported for the German version of the CTQ (Klinitzke et al. 2012). In the current study CA comprised all five scales (CTQ-tot) possibly ranging from 25-125.

<u>Urgency (affective impulsivity)</u> was measured with the German version of the Impulsive Behavior Scale, UPPS (Schmidt et al. 2008, Whiteside and Lynam 2001). UPPS urgency is self-reported scale that measures tendency for behavioral impulsivity during negative internal states regardless of consequences. It consists of twelve items that are rated with a four-point Likert scale (1-4) thus ranging from 12-48. The German version has good reliability and validity (Kämpfe and Mitte 2009).

<u>Inward anger</u> was measured with the German version of the State-Trait Aggression Inventory second edition (STAXI-2) (Rohrmann et al. 2013, Spielberger 2010). STAXI-2 has fifty-one items scored on a four-point Likert scale (1-4) and measures scales: State Anger, and trait scales, Anger Expression-In, Anger Expression-Out, Anger Control-In, and Anger Control-Out. In the current study we focused on the subscales Anger Expression-In (inward anger) and Trait Anger, which was used as a covariate in sensitivity models. Anger Expression-In measures general tendency to suppress or hold-in angry feelings instead of expressing them. It has eight items, possibly ranging from 8-32. Trait Anger measures the general tendency to experience anger, especially in negative or frustration states. It has ten items, ranging from 10-40. For the German version of the questionnaire, both subscales have good internal consistency and reliability (Etzler, Rohrmann, and Brandt 2014).

<u>Self-aggression</u> was measured using the German-developed Short Questionnaire for Assessing Factors of Aggression (Kurzfragebogen zur Erfassung von Aggressivitätsfaktoren, K-FAF; (Heubrock and Petermann 2008)). It is a self-reported questionnaire with forty-nine items with a six-point Likert scale (0-5) that measures scales: Spontaneous Aggression, Reactive Aggression, Excitability, Self-Aggression and Aggression Inhibition. Here we focused on the nine-item scale Self-aggression ranging from 0-45 that clearly delineates internally oriented aggression. We summed Spontaneous Aggression, Reactive Aggression and Excitability (thirty-three items in total, ranging from 0-165) to obtain a measure of externally oriented aggression, which we used as a covariate in sensitivity models. The internal consistency of the K-FAF is acceptable (Daig, Brähler, and Hinz 2010).

<u>Self-reported depressions severity</u> was assessed with the German version of the Beck Depression Inventory-Revision (BDI-ll; (Hautzinger et al. 1994, Beck, Steer, and Brown 1996)). The questionnaire has twenty-one items scored on a four-point Likert scale (0-3) with scores ranging from 0-63. It was shown that BDI-II has a three-factor structure, with the cognitive, somatic, and affective subscales (Vanheule et al. 2008). For sensitivity analyses we used the cognitive BDI-II subscale, comprising seven items and ranging from 0-21, which relates to the cognitive symptoms of depression that may induce negative bias.

### Data analysis

For the analysis, 2 participants were excluded due to complete missing data for the questionnaires (n= 1) and non-binary gender identity (n= 1; excluded because lack of comparability). The final sample included: SA group n= 139 (mean age [standard deviation (SD)]= 37.9 [16.3] years; 75 [54%] women); CC group n= 24 (mean age [SD]= 33.2 [12.0] years; 13 [54%] women). We compared the SA and CC groups on demographic factors using Welch’s t-test and Pearson χ2 test with Yates’ continuity correction when appropriate. **Table 1** summarizes sample demographic and clinical characteristics, and **Supplementary Table 1** summarizes SA group characteristics by study (NeST vs SUICIDE_DECIDE).

**Table 1.** Demographic and clinical characteristics of the study sample.

|  | SA group<br>n= 139 | CC group<br>n= 24 | Test statistics |
| --- | --- | --- | --- |
| Age, mean [SD] | 37.9 [16.8] | 33.2 [12.0] | <sup>A</sup> $t(40.5) = -1.7$, $p = .10$ |
| Gender, women [%] | 75 [54%] | 13 [54%] | <sup>B</sup> $\chi^2(1) < .001$, $p > .99$ |
| Primary diagnosis | | | <sup>B</sup> $\chi^2(1) = 7.7$, $p = .006$ |
| MDD | 99 [71%] | 24 [100%] |  |
| Other disorders <sup>§</sup> | 40 [29%] | / |  |
| Comorbid diagnosis <sup>§§ 1</sup> |  |  |  |
| Any | 77 [55%] | 8 [33%] | <sup>B</sup> $\chi^2(1) = 2.6$, $p = .11$ |
| PTSD | 8 [6%] | 0 [0%] | <sup>B</sup> $\chi^2(1) = 0.4$, $p = .50$ |
| Use Disorder | 41 [29%] | 4 [17%] | <sup>B</sup> $\chi^2(1) = 0.9$, $p = .34$ |
| Education <sup>2</sup> | | | <sup>B</sup> $\chi^2(2) = 5.5$, $p = .06$ |
| Elementary | 25 [18%] | 1 [4%] |  |
| High/ vocational | 95 [68%] | 17 [71%] |  |
| University | 15 [11%] | 6 [25%] |  |
| Employment | | | <sup>B</sup> $\chi^2(4) = 9.4$, $p = .051$ |
| Actively working | 56 [40%] | 11 [46%] |  |
| In education | 29 [21%] | 10 [42%] |  |
| Unemployed | 23 [16%] | 3 [12%] |  |
| Pension | 28 [20%] | 0 [0%] |  |
| Other | 3 [2%] | 0 [0%] |  |
| Kids, no [%] <sup>3</sup> | 74 [53%] | 18 [75%] | <sup>B</sup> $\chi^2(1) = 3.0$, $p = .084$ |
| Family status single, yes [%] | 92 [66%] | 18 [75%] | <sup>B</sup> $\chi^2(1) = 0.4$, $p = .54$ |

<sup>§</sup> Other disorders: Adjustment Disorder (n= 9); Autism Spectrum Disorder (n= 1); Bipolar Disorder (n= 7); Borderline Personality Disorder (n= 7); Dissociative Disorder (n= 1); Gambling Addiction Disorder (n= 1); Obsessive Compulsive Disorder (n= 1); Post Traumatic Stress Disorder (n= 3); Social Phobia (n= 1); Substance Use Disorder (n= 9).
<sup>§§</sup> Comorbid disorders included current or past.
<sup>A</sup> groups were compared using Welch two sample t-test. There were no outliers detected.
<sup>B</sup> groups were compared using $\chi^2$ test with Yates' continuity correction when appropriate.
<sup>1</sup> missing data for 1 participant in the CC group.
<sup>2</sup> missing data for 4 participants in the SA group.
<sup>3</sup> missing data for 1 participant in the SA group.
Abbreviations: CC= clinical control; MDD= Major Depressive Disorder; PTSD= Post-Traumatic Stress Disorder; SA= suicide attempt; SD= standard deviation.

Descriptive statistics for scales used in the analyses are in **Supplementary Table 2**. Scales were tested for internal consistency using Cronbach’s alpha across the sample and within the SA group. Results are described in the **Supplementary Table 3**.

For <u>Aim 1</u> we used linear regression analysis to compare CA, urgency, inward anger and self-aggression between the SA and CC groups. Based on the results of the model assumption tests, regression models were run either as linear regressions, or as robust regressions using an M estimator and bisquare weighting function. Primary model *1a* assessed effects of group. Further models were run with covariates, model *1b* with age and gender, model *1c* with age, gender, and cognitive depressive symptoms, and model *1d* with age, gender, and trait anger or externally oriented aggression for testing inward anger and self-aggression respectively. The alpha threshold was set to p< .012 to account for the four scales. In addition, we compared the childhood trauma group severity classifications based on (Häuser et al. 2011) with the German standard population classifications from (Witt et al. 2017) using χ2 tests.

For <u>Aim 2</u> within the SA group we used models to test the association of CA with urgency, inward anger, and self-aggression. Similar to Aim 1, we tested the primary model *2a* with just CTQ-tot, model *2b* with age and gender, and model *2c* with age, gender, and cognitive depressive symptoms. The alpha value was set to p< .017 to account for the three tested associations. In addition, to test the specificity of the association between the CA and inward anger versus trait anger, and self-aggression versus external aggression, estimated coefficients for each model were extracted and compared with a t-test.

Additional sensitivity analyses were performed for Aim 1 and Aim 2. Specifically, we used the Rosner test (’EnvStats’ package) to detect outliers in the behavioral variables and then re-ran the regression or mediation models excluding any detected outliers.

For analyses and graphing we used R (V4.1.3) and packages ‘sjPlot’, ‘effectsize’ and ‘car’, ‘ggplot2’, ‘cowplot’ and ‘effects’.

For <u>Aim 3</u>, we tested a theoretical parallel multiple mediation model within the SA group. In this model, CA was the predictor (X), while urgency and inward anger were modelled as parallel mediators (M1 and M2) with self-aggression as the outcome variable (Y). The parallel mediation model allows for all mediators to be examined simultaneously while accounting for the other paths and assumes that there is no causal link among the mediators (Hayes 2017). Coefficients for the two indirect, total indirect, comparison of the two indirect, direct, and total effects were computed. Analysis was done with ’lavaan (V0.6.18)’. We used only the participants with no missing variables; we evaluated the correlation matrix and calculated Mahalanobis outliers based on the 0.975 quantile of the χ2 distribution. We fitted the model using full information maximum likelihood for missing data and bootstrap estimation (1,000 samples). Standardized bootstrap estimates, bootstrap SEs, and bootstrap 95% CIs were obtained via package ‘semhelpinghands’ and bootstrap test statistics were computed (Z and p values). If a model showed any significant indirect effect (p< .05), we ran a mediation model controlling age and gender, or age, gender and cognitive depression symptoms. Supporting analyses included reliability checks (see above) and a post-hoc power analysis to inform future research. A Monte Carlo (MC) post-hoc power analysis was done by simulating datasets from the model using the standardized estimates. There were 250 replications with simulated 135 observations and models were fitted using the same settings in ‘lavaan’. Reported power is the proportion of the successful replication in which the 95% boot.CI excluded zero and 95% MC CI were computed for these proportions.

## Results

There were significant and trend-significant clinical and demographic differences between the SA and CC group, namely the primary diagnosis (% of MDD as a primary diagnosis), education, employment and presence of children (**Table 1**). There were some differences among the studies, which stemmed from the site of investigation (rural and urban areas in the NeST study and urban area of Jena in the SUICIDE_DECIDE study) (**Table S1**).

All the scales had good or excellent internal reliability, both across the whole sample and within the SA group (**Table S3**). There was one outlier in the CTQ-tot scale, while the other scales did not have outliers; therefore, additional sensitivity analyses were run only for the models that included the CTQ-tot scale. Cognitive depression symptoms (BDI-II-cog scale) did not differ between the SA and CC groups (t(36.3)= -0.3, p= .74)

### Individuals with SA and CC exhibited comparable self-reported CA severity and comparable levels of inward anger, self-aggression and urgency

On a Bonferroni corrected threshold, there were trend-significant differences between the groups for the affective impulsivity (UPPS-urgency scale) in the model controlling for age and gender (β= 3.47, 95% CI [0.61, 6.32]; p= .02, p_corr_= .067). In addition, inward anger showed trend-significant differences on an uncorrected threshold for all models, while for the CA and self-aggression there were no significant differences in any models. **Figure 1** shows the raw values and statistics for *1a* models for representation purposes while the full results are listed in **Supplementary Table S4**.

**Figure 1.**
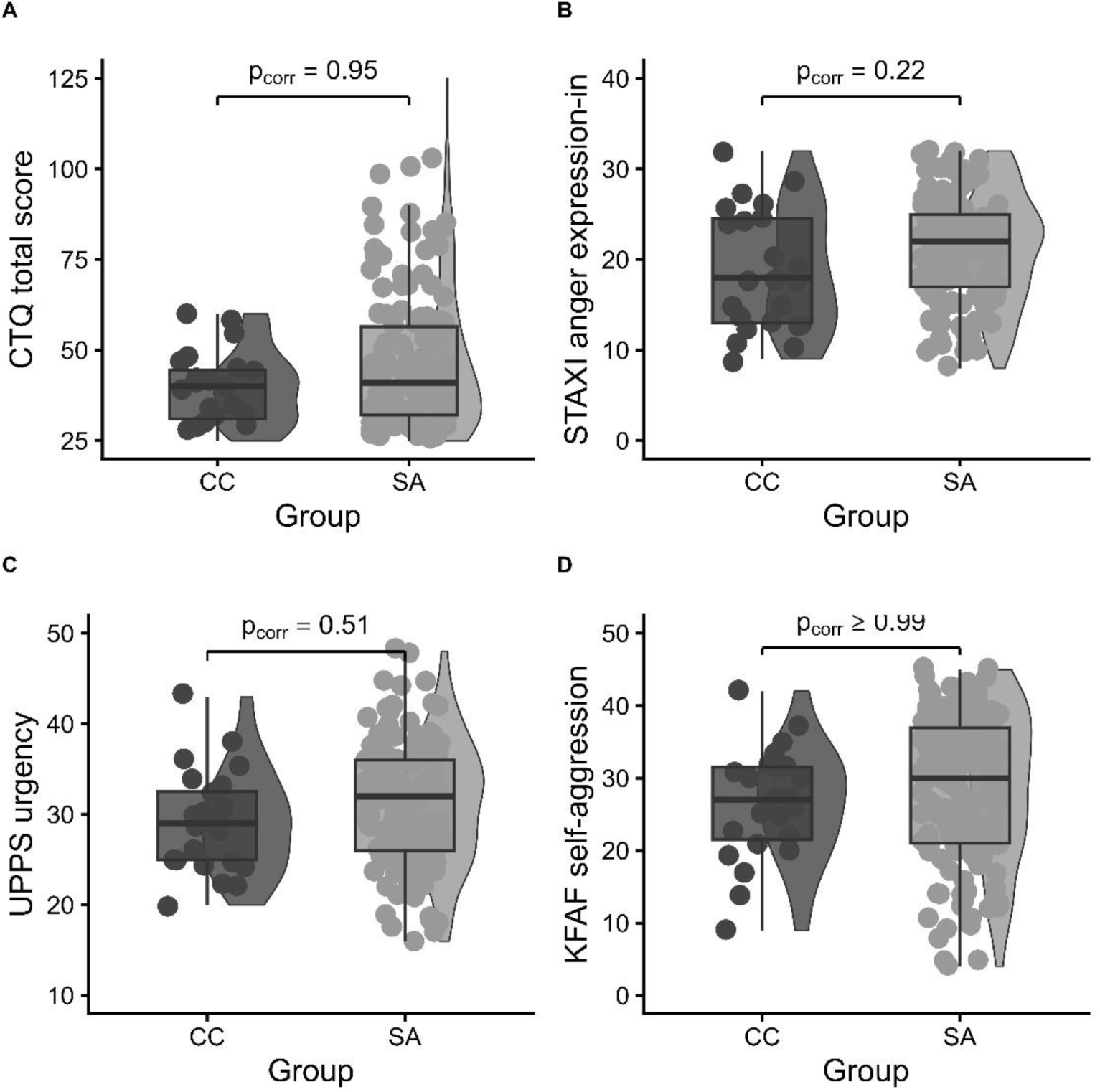
Group comparison of (A) childhood adversity measured with the CTQ-tot; (B) inward anger measured with the STAXI-2-axi; (C) impulsivity measured with the UPPS-urg; and (D) self-aggression measured with the KFAF-self. ***Note*** regression models showed that there were no significant differences between the CC and SA groups on the adjusted threshold (p< .012). Graphs show raw values for representational purposes. <u>Abbreviations</u>: CC= clinical control; CTQ-tot= total score of the Childhood Trauma Questionnaire; K-FAF= Kurzfragebogen zur Erfassung von Aggressivitätsfaktoren (Short Questionnaire for Assessing Factors of Aggression); SA= suicide attempt; STAXI-2= State-Trait Aggression Inventory second edition; UPPS= Urgency-Premeditation-Perseverance-Sensation Seeking questionnaire.

Compared with the normative German population, the SA group showed a significantly higher proportion of individuals with moderate-to-severe scores on the CTQ-SF subscales for emotional abuse, physical abuse, and emotional neglect. Detailed descriptive statistics (means [SD] and severity classifications) are presented in **Table 2**.

**Table 2.**
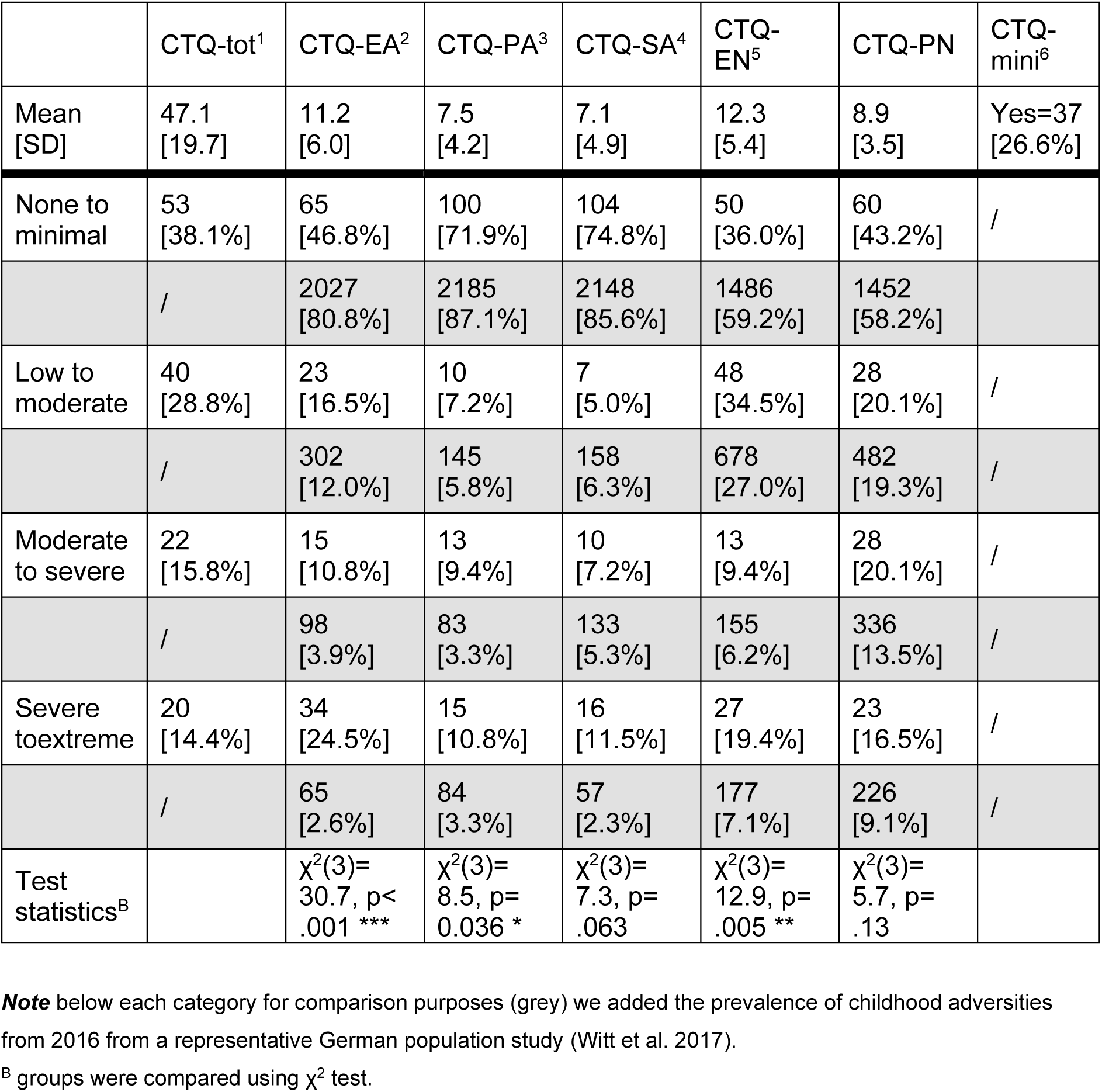
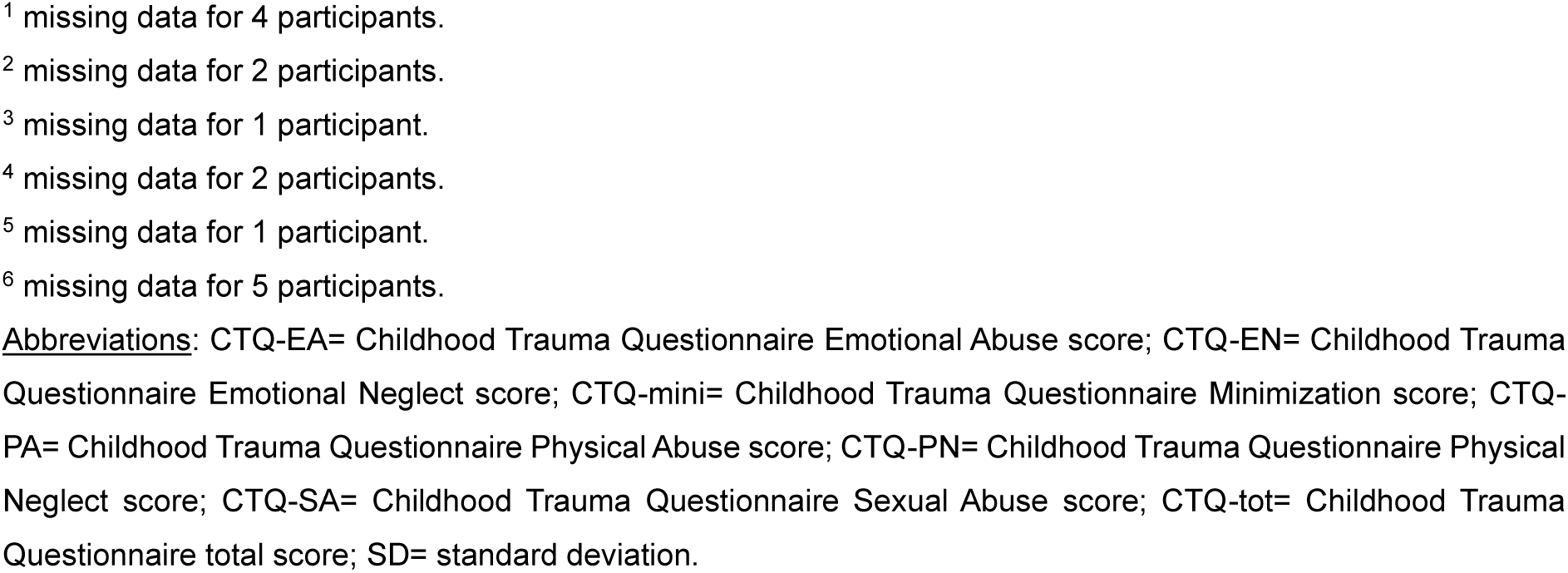
Overview of the CTQ scales and group severity classifications based on (Häuser et al. 2011) in the SA group and representative German sample.

|  | CTQ-tot<sup>1</sup> | CTQ-EA<sup>2</sup> | CTQ-PA<sup>3</sup> | CTQ-SA<sup>4</sup> | CTQ-EN<sup>5</sup> | CTQ-PN | CTQ-mini<sup>6</sup> |
| --- | --- | --- | --- | --- | --- | --- | --- |
| Mean [SD] | 47.1 [19.7] | 11.2 [6.0] | 7.5 [4.2] | 7.1 [4.9] | 12.3 [5.4] | 8.9 [3.5] | Yes=37 [26.6%] |
| None to minimal | 53 [38.1%] | 65 [46.8%] | 100 [71.9%] | 104 [74.8%] | 50 [36.0%] | 60 [43.2%] | / |
|  | / | 2027 [80.8%] | 2185 [87.1%] | 2148 [85.6%] | 1486 [59.2%] | 1452 [58.2%] |  |
| Low to moderate | 40 [28.8%] | 23 [16.5%] | 10 [7.2%] | 7 [5.0%] | 48 [34.5%] | 28 [20.1%] | / |
|  | / | 302 [12.0%] | 145 [5.8%] | 158 [6.3%] | 678 [27.0%] | 482 [19.3%] | / |
| Moderate to severe | 22 [15.8%] | 15 [10.8%] | 13 [9.4%] | 10 [7.2%] | 13 [9.4%] | 28 [20.1%] | / |
|  | / | 98 [3.9%] | 83 [3.3%] | 133 [5.3%] | 155 [6.2%] | 336 [13.5%] | / |
| Severe to extreme | 20 [14.4%] | 34 [24.5%] | 15 [10.8%] | 16 [11.5%] | 27 [19.4%] | 23 [16.5%] | / |
|  | / | 65 [2.6%] | 84 [3.3%] | 57 [2.3%] | 177 [7.1%] | 226 [9.1%] | / |
| Test statistics<sup>B</sup> | | $\chi^2(3)=30.7, p<.001$ *** | $\chi^2(3)=8.5, p=0.036$ * | $\chi^2(3)=7.3, p=.063$ | $\chi^2(3)=12.9, p=.005$ ** | $\chi^2(3)=5.7, p=.13$ | |

**Note** below each category for comparison purposes (grey) we added the prevalence of childhood adversities from 2016 from a representative German population study (Witt et al. 2017).
<sup>B</sup> groups were compared using $\chi^2$ test.
<sup>1</sup> missing data for 4 participants.
<sup>2</sup> missing data for 2 participants.
<sup>3</sup> missing data for 1 participant.
<sup>4</sup> missing data for 2 participants.
<sup>5</sup> missing data for 1 participant.
<sup>6</sup> missing data for 5 participants.
**Abbreviations:** CTQ-EA= Childhood Trauma Questionnaire Emotional Abuse score; CTQ-EN= Childhood Trauma Questionnaire Emotional Neglect score; CTQ-mini= Childhood Trauma Questionnaire Minimization score; CTQ-PA= Childhood Trauma Questionnaire Physical Abuse score; CTQ-PN= Childhood Trauma Questionnaire Physical Neglect score; CTQ-SA= Childhood Trauma Questionnaire Sexual Abuse score; CTQ-tot= Childhood Trauma Questionnaire total score; SD= standard deviation.

### Personality traits associated with self-reported CA in participants with SA

CA showed significant relation with affective impulsivity and self-aggression in the unadjusted models and adjusted for age and gender on a corrected level (p_corr_’s< .05) but these associations did not remain significant after adjustment for cognitive depression symptoms. The association with inward anger reached trend level at an uncorrected threshold (p< .07). Model comparisons suggested that CA was more strongly related to trait anger than to inward anger, and to external aggression than to self-aggression (all models p’s< .07). **Figure 2** displays the associations among the scales with complete results reported in **Supplementary Table S5**.

**Figure 2.**
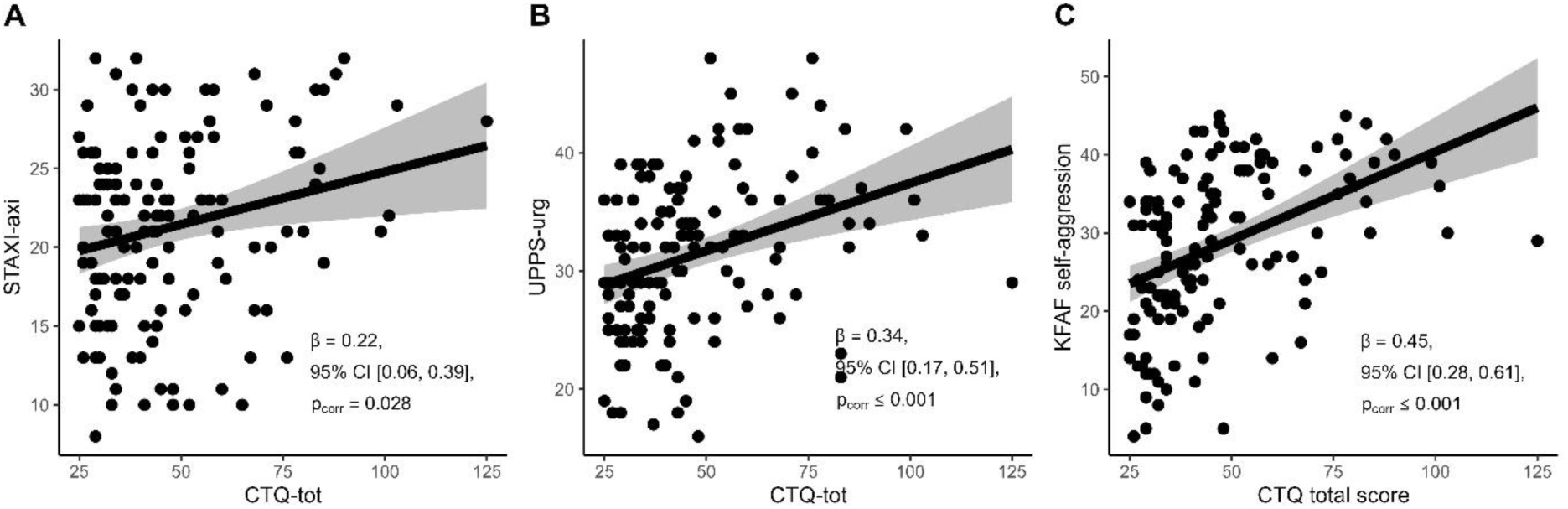
Association between childhood adversity measured with the CTQ-tot and (A) inward anger measured with the STAXI-2-axi; (B) impulsivity measured with the UPPS-urg; and (C) self-aggression measured with the KFAF-self within the SA group. ***Note*** regression models without covariates showed that there were significant associations among childhood adversity with impulsivity and self-aggression on the adjusted threshold (p< .017), but not for inward anger. Graphs show raw values for representational purposes. <u>Abbreviations</u>: CTQ-tot= total score of the Childhood Trauma Questionnaire; K-FAF= Kurzfragebogen zur Erfassung von Aggressivitätsfaktoren (Short Questionnaire for Assessing Factors of Aggression); SA= suicide attempt; STAXI-2= State-Trait Aggression Inventory second edition; UPPS= Urgency-Premeditation-Perseverance-Sensation Seeking questionnaire.

### Mediation model

Reliability analyses indicated very good internal consistency (α’s> 0.80) and there were no Mahalanobis outliers. Correlation matrix (**Supplementary table S6a**) supported the assumptions for the mediation analysis.

Parallel multiple mediation models tested whether inward anger and affective impulsivity mediated the relationship between CA and self-aggression. The model without covariates accounted for 44% of the variance in self-aggression (**Supplementary Table S6b**). Both the direct and total effects were statistically significant (p< .001). The total indirect (p< .001) was significant, as was the indirect effect of CA through affective impulsivity (p= .001), whereas the indirect effect through inward anger reached trend level (p= .070). In both paths, greater CA severity was associated with higher affective impulsivity or inward anger, which in turn was associated with greater self-aggression (**Figure 3**). A comparison of the specific indirect effects indicated that their magnitudes differed at a trend level (p= .066). The model with age and gender yielded similar results for all effects (**Supplementary Table S6c**). In contrast, the model additionally adjusting for cognitive depression symptoms showed significant effects only for the total indirect (p= .035) and total effects (p= .046) (**Supplementary Table S6d**).

**Figure 3.**
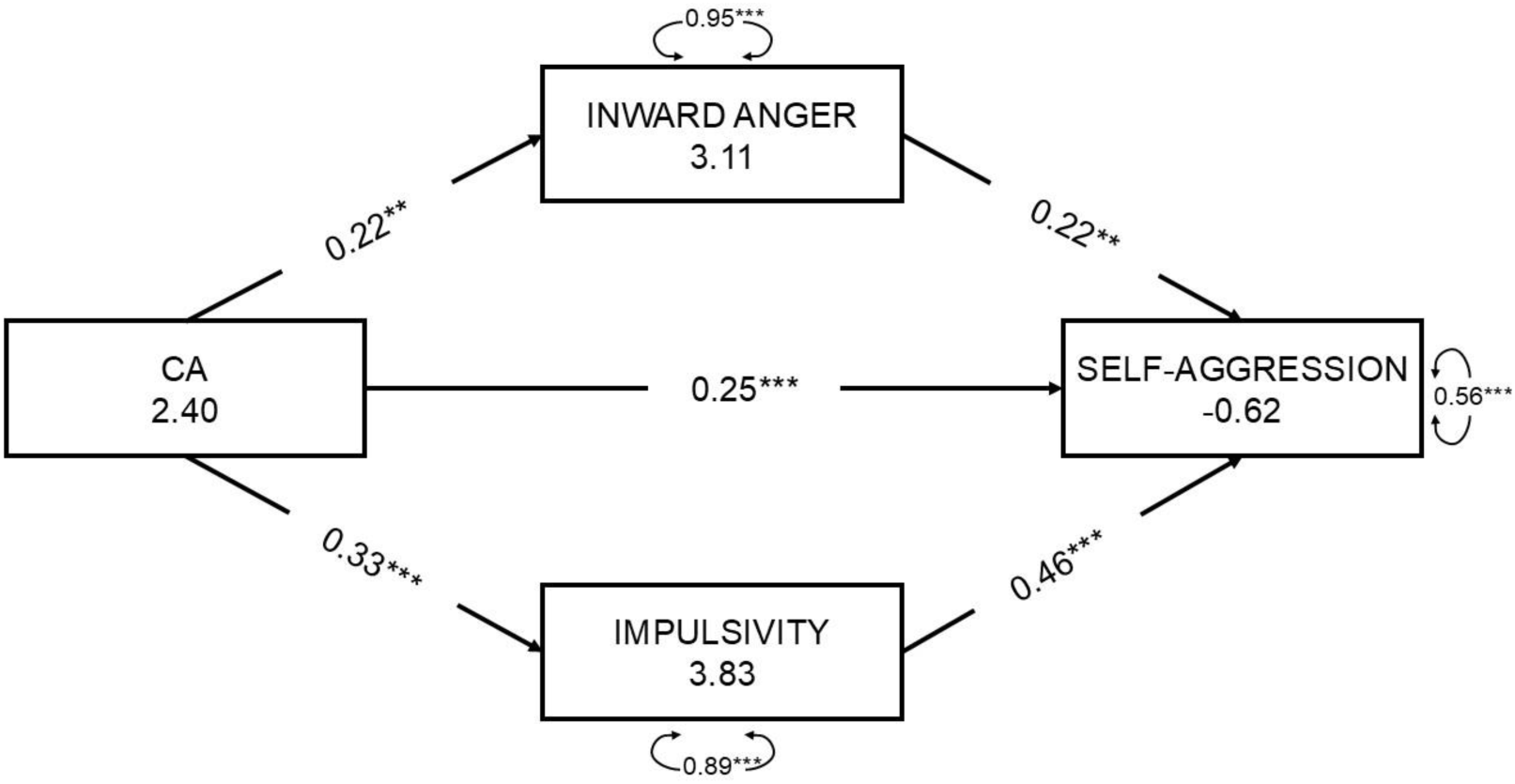
Summary of the parallel mediation model in the SA group, where the childhood adversity is the predictor variable, inward anger and impulsivity are mediators and self-aggression is the outcome variable. Model statistics indicated that the indirect path through impulsivity is significant (p= .001), while the indirect path through inward anger was not significant (p= .066). The difference between the indirect effects was not significant (p= .075). Total indirect, direct and total effects were significant (p< .001). ***Note*** model is without covariates. Full statistics for all models is in Supplementary Table S6. <u>Abbreviations</u>: CTQ-tot= total score of the Childhood Trauma Questionnaire; K-FAF= Kurzfragebogen zur Erfassung von Aggressivitätsfaktoren (Short Questionnaire for Assessing Factors of Aggression); SA= suicide attempt; STAXI-2= State-Trait Aggression Inventory second edition; UPPS= Urgency-Premeditation-Perseverance-Sensation Seeking questionnaire.

Post-hoc power Monte Carlo simulation with 250 simulations, with 1000 bootstrap resamples per fitted model indicated that the power was 0.78 95% MC CI [0.66, 0.89] for indirect effect 1; 1.0 95% MC CI [1.0, 1.0] for indirect effect 2; 1.0 95% MC CI [1.0, 1.0] for total indirect effect; and 1.0 95% MC CI [1.0, 1.0] for total effect (**Supplementary Figure 1**).

## Discussion

Epidemiological studies indicate CA as a strong distal factor for SA. Moreover, multiple theories posit that impulsivity and anger/aggression are proximal risk factors for SA. Nevertheless, previous research has used ambiguous definitions of impulsivity, anger, aggression, and suicide-related outcomes and has examined these associations across heterogeneous samples. We therefore sought to investigate these factors and their inter-relationships using well-defined constructs and questionnaires, specifically urgency (affective impulsivity during negative states), inward anger (suppression or holding-in of anger), and self-aggression (mental aggression towards oneself), in a clinically well-defined sample of individuals with a *recent* SA. In summary, the results show that:

1) there were no significant differences between SA and CC groups in investigated factors at Bonferroni-corrected significance level;
2) CA was associated with greater severity of personality factors in the SA group, though associations with overall trait anger and external aggression were also observed;
3) there were indirect effects of CA through affective impulsivity on self-aggression, which were somewhat stronger compared with the indirect effect of CA through inward anger.

In support of CA being prevalent in persons with SA, the SA group showed a higher proportion of individuals in the moderate and severe categories across types of adversities measured with the CTQ-SF, compared to a representative German sample (Witt et al. 2017, Witt et al. 2018). Emotional abuse and emotional neglect were the most frequently endorsed CA categories in the SA group. Multiple types of childhood trauma have been connected with increased risk for SA, while some studies highlight emotional abuse as the specifically relevant form (De Araújo and Lara 2016, Liu et al. 2017). Emotional abuse may represent a specifically salient type of CA, as it may promote maladaptive negative affect regulation in interpersonal situations, for example through suppression (Zhou and Zhen 2022) and internally oriented anger, as reported in non-SA populations (Gardner, Thomas, and Erskine 2019), a pattern consistent with the elevated inward anger observed in individuals with CA in our SA group (though at trend level).

Importantly, SA group did not significantly differ from CC in overall CA severity (CTQ-tot score), suggesting that CA may be more broadly associated with psychiatric vulnerability rather than being a specific risk factor for SA. This is consistent with a literature linking CA to a wide range of psychiatric disorders (Hogg et al. 2023, Kessler et al. 2010), particularly depressive disorders (King 2021), and supports the view that CA may represent a transdiagnostic risk factor rather than a pathway specific to suicidal behavior. Notably, the SA group showed greater variance in overall CA severity compared to the CC group (Figure 1A), which may reflect meaningful heterogeneity within the SA population itself. Rather than representing a homogeneous group uniformly characterized by high CA burden, individuals with SA may constitute distinct subgroups, who may differ in their underlying pathways to suicidal behavior. Future studies with larger sample sizes could identify CA-based subgroups within SA samples, for instance through cluster analytic approaches, to better characterize which individuals with SA are specifically characterized by CA burden and whether these subgroups differ in the psychological factors examined here, such as impulsivity, inward anger, and self-aggression.

Moreover, for personality factors, SA and CC group differed only marginally in affective impulsivity (urgency) suggesting inward anger and self-aggression are comparable across clinical samples. An older study across clinical samples in Germany described that ‘endogenous depression’ (ICD-9) had higher levels of inward anger-expression compared to psychosomatic and anxiety disorders (Müller et al. 2001) which may explain why we did not find significant differences as both groups had primary diagnoses of depression (for SA group mostly and for CC group exclusively). Other research, such as a meta-analysis (Beach, Gissandaner, and Schmidt 2022) also suggests that personality factors like trait impulsivity may be related to underlying psychopathology and are not necessarily the best factors for distinguishing or predicting SA.

As hypothesized, within the SA group, CA severity and personality factors were strongly associated with each other (**Figure 2**; **Table S5**). Nevertheless, these associations were not specific to internal facets as stronger associations were observed for external facets, namely trait anger and external aggression. Several explanations are possible. First, the questionnaires used may not adequately capture the distinction between “internally” and “externally” oriented anger and aggression in individuals with SA. Second, a latent general factor of anger and aggression may underly both internal and external facets, rendering their differentiation difficult at the questionnaires level. Third, the persons with SA may have maladaptive cognitive schemas that blur the boundary between internally and externally directed reactivity, such that anger and aggression are experienced and expressed in a less differentiated way than in non-SA populations. For example, previous studies in German samples showed a strong association among self-aggression and external forms of aggression both in persons with and without psychopathology (Otte et al. 2019) indicating a strong overlap among the forms and a possible general form of aggression that may be related to SA. This overlap may also reflect a fundamental limitation at the measurement level, as currently available questionnaires may not sufficiently differentiate between internally and externally directed aggression as distinct constructs, particularly in clinical populations where these boundaries may be inherently blurred.

The theoretical parallel mediation models showed significant indirect effects (**Figure 3**), with significant indirect effects of CA through affective impulsivity and trend-significant though inward anger. The findings are in line with previous studies that also showed mediation effects of various measures of impulsivity from CA to suicidal behavior (Pérez-Balaguer et al. 2022, Yarar, Bulut, and Demirbaş 2023, Dal Santo et al. 2020) and mediating effect of anger from CA to suicide behavior in adolescents (Sigfusdottir et al. 2013) and future attempts in adults (Rabbany et al. 2023). An older study in persons with a lifetime suicide attempt found association among impulsivity (measured with Temperament and character inventory) and facets of anger (measured with STAXI) on general and self-aggression (measured with Fragebogen zur Erfassung von Aggressivitätsfaktoren-FAF) (Giegling et al. 2009). In addition a meta-analysis reported association among trait impulsivity factors and facets of aggression (verbal, physical and general) across studies in clinical and non-clinical populations, suggesting trait impulsivity is a robust factor of overall aggression (Bresin 2019).

Overall, the theoretical model requires further conformation, as the observed effects may relate epiphenomena of underlying depressive psychopathology rather than independent causal pathways. This is suggested by the pattern of sensitivity analyses, in which the inclusion of cognitive depressive symptoms as a covariate altered the variance explained in the regression and mediation models. In support, meta-analysis on impulsivity and aggression also proposed that the association of these factors with suicidality (encompassing suicidal ideation, suicide attempts, death by suicide, and related outcomes) are positive and weak and are part of underlying psychopathology (Moore et al. 2022). Taken together, these findings highlight the need for longitudinal designs that can better disentangle trait-level vulnerability factors from state-dependent depressive symptoms in the pathway from CA to suicidal behavior.

### Limitations

The study was cross-sectional, and the associations may relate to current levels of psychopathology. In addition, the assessment of self-aggression via K-FAF was highly correlated with depression (BDI-II), which may reflect a methodological limitation rather than a true conceptual overlap. Self-aggression, as a relatively underexplored construct in the context of suicidal behavior, may benefit from more refined assessment tools that are specifically designed to capture aggression directed toward oneself independently of current mood state or depressive symptom burden. The development or adaptation of such instruments would allow future studies to disentangle self-aggression as a stable trait-like vulnerability factor from state-dependent fluctuations associated with acute depressive episodes, thereby improving both the construct validity and the clinical utility of this measure in suicide research. The CC group were smaller and acquired only in one study. Mediation models were theoretical in nature. Even though the participants with SA had their most recent attempt <12 months, the tested associations were conducted after the most recent SA and can thus not show any predictive value for future SA.

### Outlook

Population studies indicate high prevalence of CA, which carries high attributable proportion for risk for SB (Grummitt et al. 2024). However, as our findings suggest, the SA group itself is likely heterogeneous with respect to CA burden, with CA representing a salient risk pathway for some individuals but not others. Our results, as in line with multiple theoretical frameworks of suicidal crisis, point out that CA may act through several pathways or proximal factors in a non-specific manner (Hostinar et al. 2023). Moreover, individual risk factor measures have shown limited predictive power (Ribeiro et al. 2016). We propose that persistent maladaptive coping strategies for regulating negative affect, for example inward anger during interpersonal conflicts, may contribute to SA only as part of a specific bio-socio-psychological pattern. Additionally, most studies (including ours) focus only on risk factors, while the simultaneous assessment of protective factors or their absence may deepen our understanding of suicidal crises and suicidal behavior (McClatchey et al. 2019). Future research should thus orient into measuring multiple biological, socio-environmental, cognitive and affective domains simultaneously and longitudinally to reveal bio-socio-psychological patterns i) across persons thus aiding development of short interventions that can be broadly disseminated (Bahlmann et al. 2022) or ii) within persons across time, supporting the development of digital personalized approaches (Braciszewski 2021).

### Conclusions

We highlight the prevalence of different types of CA measured with CTQ-SF in a German sample of participants with *recent* SA. Moreover, we report higher levels of urgency in participants with recent SA compared with CC group. Within SA group higher CA was related to an overall higher severity of proximal risk factors, however the association of CA with anger and aggression subscales was unspecific to internal vs external indicating the need for better definitions and questionnaires developed specifically for individuals with SA. Urgency and inward anger were similarly mediating association among CA and self-aggression indicating that both factors may contribute independently to the heightened risk for SA in high-risk situations. Our results suggest that these factors may be relevant for consideration in psychobehavioral therapy for individuals at risk for SA and those with broader psychopathology.

## Data Availability

All data produced in the present study are available upon reasonable request to the authors. Data sharing must pertain to the requirements by the local ethic comittee.

## Authors contributions

LC: Conceptualization; Data Curation; Formal analysis; Supervision; Writing - Original Draft

JM: Investigation; Data Curation; Formal analysis; Writing - Original Draft

AZ: Investigation; Data Curation; Writing - Review & Editing

LB: Investigation; Data Curation; Writing - Review & Editing

BB: Formal analysis; Writing - Review & Editing

MW: Writing - Review & Editing

FJ: Funding acquisition; Investigation; Writing - Review & Editing

GW: Funding acquisition; Conceptualization; Investigation; Data Curation; Supervision; Writing - Original Draft

## Declaration of generative AI and AI-assisted technologies in the manuscript preparation process

During the preparation of this work the authors used Gemini 3 Flash and Nature Research Assistant to improve grammar and clarity of paragraphs. After using these tools, the authors reviewed and edited the content as needed and took full responsibility for the content of the published article.

## Acknowledgement

We thank our participants for their participation in our studies. We thank researchers and staff that participated in the data collection (Ms. Marlehn Lübbert, Ms. Johanna Walther, Ms. Anna Karoline Seiffert and Ms. Anna Bollmann).

