## Supplementary Material for "Personality factors and childhood adversity in psychiatric patients with and without recent suicide attempts: a cross-sectional study"

**Supplementary Table S1** Demographic and clinical characteristics of the SA group by study.

|  | NEST study  n= 99 | BADO study n= 40 | Test statistics |
| --- | --- | --- | --- |
| Age, mean [SD] | 40.9 [17.8] | 30.6 [11.3] | ^A^ t(111.9)= -4.1, p< .001 |
| Gender, women [%] | 47 [47%) | 28 [70%] | ^B^ χ^2^(1)= 4.9, p= .026 |
| Primary diagnosis  MDD  Other disorders ^§^ | 63 [64%]  36 [36%] | 36 [90%]  4 [10%] | ^B^ χ^2^(1)= 8.4, p= .004 |
| Comorbid diagnosis ^§§^  Any  PTSD  Use Disorder | 51 [51%]  4 [4%]  25 [25%] | 26 [65%]  4 [10%]  16 [40%] | ^B^ χ^2^(1)= 1.6, p= .21  ^B^ χ^2^(1)= 0.9, p= .33  ^B^ χ^2^(1)= 3.0, p= .084 |
| Education ^1^  Elementary  High/ vocational  University | 23 [23%]  67 [68%]  7 [7%] | 2 [5%]  28 [70%]  8 [20%] | ^B^ χ^2^(2)= 9.8, p= .007 |
| Employment  Actively working  In education  Unemployed  Pension  Other | 37 [37%]  17 [17%]  16 [16%]  27 [27%]  2 [2%] | 19 [47%]  12 [30%]  7 [18%]  1 [2%]  1 [3%] | ^B^ χ^2^(4)= 11.7, p= .020 |
| Kids, no [%] ^2^ | 47 [47%] | 27 [67%] | ^B^ χ^2^(1)= 3.6, p= .057 |
| Family status single, yes [%] | 67 [68%] | 25 [62%] | ^B^ χ^2^(1)= 0.1, p= .70 |

^§^ Other disorders (NEST/ BADO): Adjustment Disorder (n= 8/ 1); Autism (n= 1/ 0); Bipolar Disorder (n= 7/ 0); Borderline Personality Disorder (n= 7/ 0); Dissociative Disorder (n= 1/ 0); Gambling Addiction Disorder (n= 1/ 0); Obsessive Compulsive Disorder (n= 1/ 0); Post Traumatic Stress Disorder (n= 1/ 2); Social Phobia (n= 0/ 1); Substance Use Disorder (n= 9/ 0).

^§§^ Comorbid disorders included current or past.

^1^ Missing data for 2 participants in the NEST study and 2 participants in the BADO study.

^2^ Missing data for 1 participant in the NEST study.

^A^ groups were compared using Welch two sample t-test. There were no outliers.

^B^ groups were compared χ2 test with Yates' continuity correction when appropriate.

Abbreviations: BADO= The choice of a violent suicidal means: a MRI study with computational modeling of decision-making study; MDD= Major Depressive Disorder; NEST= Network for suicide prevention in Thuringia study; PTSD= Post-Traumatic Stress Disorder; SA= suicide attempt; SD= standard deviation.

**Supplementary Table S2**. Descriptive values for psychometric scales.

|  | SA group | CC group |
| --- | --- | --- |
| BDI-II-cog ^1^ | 9.78 [5.81] (0-21) | 9.43 [4.37] (2-17) |
| CTQ-tot ^2^ | 47.1 [19.7] (25-125) | 39.4 [9.84] (25-60) |
| STAXI-2 anger expression-in ^3^ | 21.2 [5.80] (8-32) | 18.7 [6.67] (9-32) |
| STAXI-2 trait anger ^3^ | 22.0 [7.15] (11-40) | 21.2 [4.85] (13-32) |
| UPPS urgency ^4^ | 31.4 [6.74] (16-48) | 29.2 [5.66] (20-43) |
| K-FAF self-aggression ^5^ | 28.5 [10.1] (4-45) | 26.4 [7.72] (9-42) |
| K-FAF external aggression ^§ 5^ | 52.9 [27.8] (5-132) | 46.0 [20.8] (12-91) |

***Note***: data is presented as mean [SD] (minimum-maximum). Rosner test detected one outlier in the CTQ-tot (in the SA group); other scales did not have outliers.

^§^ represents sum score of K-FAF spontaneous aggression, reactive aggression and excitability aggression.

^1^ missing data for 1 participant in the CC group.

^2^ missing data for 4 participants in the SA group and 1 participant in the CC group.

^3^ missing data for 1 participant in the SA group and 1 participant in the CC group.

^4^ missing data for 3 participants in the SA group and 1 participant in the CC group.

^5^ missing data for 1 participant in the CC group.

Abbreviations: BDI-II-cog= cognitive subscore of the Beck Depression Inventory- Revision; CC= clinical control; CTQ-tot= total score of the Childhood Trauma Questionnaire; K-FAF= Kurzfragebogen zur Erfassung von Aggressivitätsfaktoren (Short Questionnaire for Assessing Factors of Aggression); SA= suicide attempt; SD= standard deviation; STAXI-2= State-Trait Aggression Inventory second edition; UPPS= Urgency-Premeditation-Perseverance-Sensation Seeking questionnaire.

**Supplementary Table S3**. Internal consistency statistics for the psychometric scales across the whole sample and within the SA group.

|  | Number of items | Whole sample | SA group |
| --- | --- | --- | --- |
| BDI-II-cog ^1^ | 7 | α= 0.88 | α= 0.90 |
| CTQ-tot ^2^ | 25 | α= 0.94 | α= 0.94 |
| STAXI-2 anger expression-in ^3^ | 8 | α= 0.89 | α= 0.88 |
| STAXI-2 trait anger ^3^ | 10 | α= 0.92 | α= 0.93 |
| UPPS urgency ^4^ | 12 | α= 0.83 | α= 0.84 |
| K-FAF self-aggression ^5^ | 9 | α= 0.83 | α= 0.85 |
| K-FAF external aggression ^§ 5^ | 33 | α= 0.92 | α= 0.93 |

***Note***: Cronbach’s α based upon the covariances.

^§^ represents sum score of K-FAF spontaneous aggression, reactive aggression and excitability aggression with Cronbach’s α= 0.81, 0.83 and 0.90 respectively.

^1^ missing data for 1 participant in the CC group.

^2^ missing data for 4 participants in the SA group and 1 participant in the CC group.

^3^ missing data for 1 participant in the SA group and 1 participant in the CC group.

^4^ missing data for 3 participants in the SA group and 1 participant in the CC group.

^5^ missing data for 1 participant in the CC group.

Abbreviations: BDI-II-cog= cognitive subscore of the Beck Depression Inventory- Revision; CTQ-tot= total score of the Childhood Trauma Questionnaire; K-FAF= Kurzfragebogen zur Erfassung von Aggressivitätsfaktoren (Short Questionnaire for Assessing Factors of Aggression); SA= suicide attempt; SD= standard deviation; STAXI-2= State-Trait Aggression Inventory second edition; UPPS= Urgency-Premeditation-Perseverance-Sensation Seeking questionnaire.

**Supplementary Table S4.** Summary of regression models testing the difference between the SA and CC groups for childhood adversity, inward anger, impulsivity, and self-aggression. For brevity, we noted statistics for the group variable.

|  | *Model 1a* | *Model 1b* | *Model 1c* | *Model 1c* |
| --- | --- | --- | --- | --- |
| ***CTQ-tot*** | | | | |
| Group^§^ | B= 4.1, 95% CI [-2.94, 11.25]; β= 0.22, 95% CI [-0.16, 0.60]; p= .24, p_corr_= .95 | B= 4.9, 95% CI [-1.63, 11.41]; β= 0.26, 95% CI [-0.09, 0.61]; p= .13, p_corr_= .52 | B= 3.7, 95% CI [-2.48, 9.82]; β= 0.20, 95% CI [-0.13, 0.53]; p= .23, p_corr_= .92 |  |
| *Models’ statistics* | Residual st.error (156)= 15.4 | Residual st.error (154)= 13.3 | Residual st.error (153)= 12.6 |  |
| *Models’ metrics* | AIC_c_=1382.1 (0.06)  Sigma= 18.95 | AIC_c_=1372.5 (0.02)  Sigma= 18.25 | AIC_c_=1357.8 (0.03)  Sigma= 17.36 |  |
| ***STAXI-2-anger expression-in*** | | | | |
| Group^§^ | B= 2.8, 95% CI [-0.02, 5.60]; β= 0.47, 95% CI [-0.01, 0.94]; p= .055, p_corr_= .22 | B= 2.6, 95% CI [-0.04, 5.29]; β= 0.44, 95% CI [-0.01, 0.88]; p= .053, p_corr_= .21 | B= 2.3, 95% CI [-0.29, 4.89]; β= 0.38, 95% CI [-0.05, 0.82]; p= .081, p_corr_= .33 | B= 2.2, 95% CI [-0.39, 4.76]; β= 0.36, 95% CI [-0.7, 0.79]; p= .096 |
| *Models’ statistics* | Residual st.error (158)= 6.5 | R^2^= 0.03 Adj. R^2^= 0.01, p= .17 | R^2^= 0.09 Adj. R^2^= 0.07, p= .004 | R^2^= 0.11 Adj. R^2^= 0.09, p= .002 |
| *Models’ metrics* | AIC_c_= 1028.9 (0.5)  Sigma= 5.95 | AIC_c_= 1031.3 (0.5)  Sigma= 5.95 | AIC_c_= 1023.1 (0.5)  Sigma= 5.78 | AIC_c_= 1020.9 (0.5)  Sigma= 5.72 |
| ***UPPS-urgency*** | | | | |
| Group^§^ | B= 2.3, 95% CI [-0.66, 5.21]; β= 0.34, 95% CI [-0.10, 0.79]; p= .13, p_corr_= .51 | B= 3.5, 95% CI [0.63, 6.30]; β= 3.47, 95% CI [0.61, 6.32]; p= .02, p_corr_= .067 | B= 2.6, 95% CI [-0.05, 5.18]; β= 2.57, [-0.05, 5.19]; p= .055, p_corr_= .22 |  |
| *Models’ statistics* | R^2^= 0.01, Adj. R^2^= 0.008, p= .13 | Residual st.error (155)= 5.97 | R^2^= 0.25, Adj. R^2^= 0.23, p< .001 |  |
| *Models’ metrics* | AIC_c_= 1055.5 (0.5)  Sigma= 6.60 | AIC_c_= 1034.2 (0.5)  Sigma= 6.13 | AIC_c_= 1019.2 (0.5)  Sigma= 5.83 |  |
| ***KFAF-self aggression*** | | | | |
| Group^§^ | B= 2.3, 95% CI [-2.30, 6.85]; β= 0.23, 95% CI [-0.24, 0.70]; p= .32, p_corr_> .99 | B= 3.2, 95% CI [-0.60, 7.04]; β= 0.33, 95% CI [-0.06, 0.72]; p= .098, p_corr_= .39 | B= 2.0, 95% CI -0.95, 4.88]; β= 0.20, 95% CI [-0.10, 0.50]; p= .18, p_corr_= .74 | B= 1.5, 95% CI [-1.25, 4.22]; β= 0.15, 95% CI [-0.13, 0.43]; p= .30 |
| *Models’ statistics* | Residual st.error (160)= 10.9 | R^2^= 0.25 Adj. R^2^= 0.24, p< .001 | R^2^= 0.57 Adj. R^2^= 0.56, p< .001 | Residual st.error (156)= 5.9 |
| *Models’ metrics* | AIC_c_= 1203.7 (0.5)  Sigma= 9.81 | AIC_c_= 1161.4 (0.5)  Sigma= 8.55 | AIC_c_= 1073.6 (0.5)  Sigma= 6.50 | AIC_c_= 1044.5 (0.5)  Sigma= 5.92 |

***Note*** based on the results of model assumption tests, regression models were run either as linear regressions, or as robust regressions using an M estimator and bisquare weighting function. Models were run as simple models comparing group effects (*1a*), models controlled for age and gender (*1b*), age, gender and cognitive depressive symptoms (*1c*) or age, gender, and temperament anger or externally oriented aggression for testing inward anger and self-aggression respectively (*1d*).

The alpha value was set to p< .012 to account for testing four scales.

^§^ reference group is the CC group.

^1^ CTQ-tot had one outlier; therefore, we did a sensitivity analysis without that participant. The statistics was almost the same, here in brief: *Model 1a*, β= -0.23, 95% CI [-0.17, 0.64], p= .24; *Model 1b*, β= 0.28, 95% CI [-0.09, 0.64], p= .13; *Model 1c*, β= 0.20, 95% CI [-0.14, 0.55], p= .23.

Abbreviations: AIC_C_= Akaike Information Criterion, with a correction for small sample sizes; BDI-II-cog= cognitive subscore of the Beck Depression Inventory- Revision; CC= clinical control; CTQ-tot= total score of the Childhood Trauma Questionnaire; K-FAF= Kurzfragebogen zur Erfassung von Aggressivitätsfaktoren (Short Questionnaire for Assessing Factors of Aggression); SA= suicide attempt; STAXI-2= State-Trait Aggression Inventory second edition; st.error= standard error; UPPS= Urgency-Premeditation-Perseverance-Sensation Seeking questionnaire; p_corr_= corrected p value using Bonferroni correction.

**Supplementary Table S5.** Summary of regression models of the association of childhood adversity with inward anger, impulsivity and self-aggression within the SA group. For brevity, we noted statistics for the CA variable.

|  | *Model 2a* | *Model 2b* | *Model 2c* |  |  |  |
| --- | --- | --- | --- | --- | --- | --- |
| ***STAXI-2 anger expression-in*** | | | |  |  |  |
| CTQ-tot | B= 0.07, 95% CI [0.02, 0.12]; β= 0.22, 95% CI [0.06, 0.39]; p= .009, p_corr_= .028 | B= 0.06, 95% CI [0.01, 0.12]; β= 0.21, 95% CI [0.03, 0.39]; p= .02, p_corr_= .061 | B= 0.04, 95% CI [-0.01, 0.10]; β= 0.14, 95% CI [-0.04, 0.33]; p= .13, p_corr_= .40 |  |  |  |
| *Models’ statistics* | R^2^= 0.05, Adj. R^2^= 0.04, p= .009 | R^2^= 0.05, Adj. R^2^= 0.03, p= .074 | R^2^= 0.09, Adj. R^2^= 0.06, p= .016 |  |  |  |
| *Models’ metrics* | AIC_c_= 846.6 (0.8)  Sigma= 5.74 | AIC_c_= 850.6 (0.6)  Sigma= 5.78 | AIC_c_= 847.4 (0.5)  Sigma= 5.69 |  |  |  |
| *Coeff. comparison*^§^ | Z= -2.48, p= .013 | Z= -2.05, p= .040 | Z= -1.82, p= .068 |  |  |  |
| ***UPPS-urg*** | | | |  |  |  |
| CTQ-tot | B= 0.1, 95% CI [0.06, 0.17]; β= 0.34, 95% CI [0.17, 0.51]; p< .001, p_corr_< .001 | B= 0.08, 95% CI [0.03, 0.14]; β= 0.25, 95% CI [0.08, 0.42]; p= .005, p_corr_= .015 | B= 0.05, 95% CI [-0.002, 0.11]; β= 0.16, 95% CI [-0.01, 0.32]; p= .059, p_corr_= .18 |  |  |  |
| *Models’ statistics* | Residual st.error (130)= 6.6 | Residual st.error (128)= 5.9 | R^2^= 0.29, Adj. R^2^= 0.26, p< .001 |  |  |  |
| *Models’ metrics* | AIC_c_= 869.2 (0.5)  Sigma= 6.41 | AIC_c_= 856.9 (0.5)  Sigma= 6.06 | AIC_c_= 846.9 (0.5)  Sigma= 5.81 |  |  |  |
| ***KFAF-self aggression*** | | | |  |  | Z= -2.86, p= .004 |
| CTQ-tot | B= 0.2, 95% CI [0.15, 0.31]; β= 0.45, 95% CI [0.28, 0.61]; p< .001, p_corr_< .001 | B= 0.2, 95% CI [0.08, 0.24]; β= 0.32, 95% CI [0.17, 0.46]; p< .001, p_corr_< .001 | B= 0.06, 95% CI [-0.003, 0.12]; β= 0.11, 95% CI [-0.01, 0.23]; p= .066, p_corr_= .20 |  |  |  |
| *Models’ statistics* | Residual st.error (133)= 9.9 | R^2^= 0.35, Adj. R^2^= 0.33, p< .001 | Residual st.error (130)= 6.5 |  |  |  |
| *Models’ metrics* | AIC_c_= 982.0 (0.5)  Sigma= 9.05 | AIC_c_= 958.2 (0.5)  Sigma= 8.22 | AIC_c_= 877.3 (0.5)  Sigma= 6.06 |  |  |  |
| *Coeff. comparison*^§^ | Z= -2.95, p= .003 | Z= -3.48, p< .001 | Z= -3.46, p< .001 |  |  |  |

***Note*** based on the results of model assumption tests, regression models were run either as linear regressions, or as robust regressions using an M estimator and bisquare weighting function. Models were run as simple models (*2a*), models controlled for age and gender (*2b*), or age, gender and cognitive depressive symptoms (*2c*). The alpha value was set to p< .017 to account for testing three scales.

^§^ Coefficients were compared using a t-test. Reference models were with STAXI-2-axi compared with STAXI-2-trait, and KFAF-self aggression compared with KFAF-external. The results showed that the CTQ-tot is significantly more predictive of trait anger compared to inward anger, and external aggression compared to self-aggression.

^1^ CTQ-tot had one outlier; therefore, we did a sensitivity analysis without that participant. The statistics was almost the same, here in brief:

-STAXI-2-axi *Model 1a*, β= 0.20, 95% CI [0.03, 0.37], p= .019, coeff.comp p= .002; *Model 1b*, β= 0.19, 95% CI [0.01, 0.37], p= .04, coeff.comp p= .009; *Model 1c*, β= 0.11 95% CI [-0.08, 0.30], p= .25, coeff.comp p= .016.

-UPPS-urg *Model 1a*, β= 0.37, 95% CI [0.21, 0.53], p< .001; *Model 1b*, β= 0.28 95% CI [0.11, 0.45], p= .001; *Model 1c*, β= 0.18 95% CI [0.02, 0.35], p= .032.

-KFAF-self *Model 1a*, β= 0.47, 95% CI [0.31, 0.63], p< .001, coeff.comp p< .001; *Model 1b*, β= 0.34, 95% CI [0.19, 0.49], p< .001, coeff.comp p< .001; *Model 1c*, β= 0.13, 95% CI [0.01, 0.25], p= .04, coeff.comp p< .001.

Abbreviations: coeff.= coefficient; CTQ-tot= total score of the Childhood Trauma Questionnaire; K-FAF= Kurzfragebogen zur Erfassung von Aggressivitätsfaktoren (Short Questionnaire for Assessing Factors of Aggression); STAXI-2= State-Trait Aggression Inventory second edition; st.error= standard error; UPPS= Urgency-Premeditation-Perseverance-Sensation Seeking questionnaire; p_corr_= corrected p value using Bonferroni correction.

**Supplementary Table S6.** Parallel mediation model. Testing effects of childhood adversity on self-aggression through inward anger and impulsivity.

**Supplementary Table S6a.** Means, standard deviations, and correlations with confidence intervals.

| Variable | M | SD | 1 | 2 | 3 |
| --- | --- | --- | --- | --- | --- |
| CTQ-tot | 47.07 | 19.70 |  |  |  |
| STAXI-2-axi | 21.21 | 5.80 | .22** |  |  |
|  |  |  | [.06, .38] |  |  |
| UPPS-urg | 31.45 | 6.74 | .34** | .32** |  |
|  |  |  | [.17, .48] | [.15, .46] |  |
| KFAF-self | 28.45 | 10.10 | .44** | .40** | .59** |
|  |  |  | [.30, .57] | [.25, .53] | [.47, .69] |

***Note*** M and SD are used to represent mean and standard deviation, respectively. Values in square brackets indicate the 95% confidence interval for each correlation. * indicates p < .05. ** indicates p < .01.

***Note*** There were no Mahalanobis outliers based on the 0.975 quantile of the chi-square distribution

**Supplementary Table S6b.** Model coefficients assessing inward anger and impulsivity as parallel mediators from childhood adversity to self-aggression. Model is without covariates.

| **Outcome / Path** | **β** | **SE (boot)** | **Z (boot)** | **p (boot)** | **95% CI (boot lower, upper)** | **R²** |
| --- | --- | --- | --- | --- | --- | --- |
| STAXI-2-axi: intercept (~1) | 3.11 | 0.29 | 10.59 | < .001 | 2.58, 3.77 | 0.05 |
| STAXI-2-axi ~ CTQ-tot (a1) | 0.22 | 0.08 | 2.99 | .003 | 0.07, 0.36 |  |
| UPPS-urg: intercept (~1) | 3.83 | 0.37 | 10.26 | < .001 | 3.13, 4.57 | 0.11 |
| UPPS-urg ~ CTQ-tot (a2) | 0.34 | 0.08 | 4.24 | < .001 | 0.19, 0.50 |  |
| KFAF-self: intercept (~1) | −0.62 | 0.38 | −1.62 | .105 | −1.35, 0.11 | 0.44 |
| KFAF-self~ STAXI-2-axi (b1) | 0.22 | 0.08 | 2.76 | .006 | 0.06, 0.37 |  |
| KFAF-self ~ UPPS-urg (b2) | 0.46 | 0.07 | 6.56 | < .001 | 0.32, 0.59 |  |
| KFAF-self~CTQ-tot (directc) | 0.25 | 0.07 | 3.79 | < .001 | 0.12, 0.39 |  |

| **Effect** | **β** | **SE (boot)** | **Z (boot)** | **p (boot)** | **95% CI (boot lower, upper)** |
| --- | --- | --- | --- | --- | --- |
| Direct (c) | 0.25 | 0.07 | 3.79 | < .001 | 0.12, 0.39 |
| Indirect 1 (a1*b1) | 0.05 | 0.03 | 1.81 | .070 | −0.00, 0.11 |
| Indirect 2 (a2*b2) | 0.15 | 0.05 | 3.42 | .001 | 0.07, 0.25 |
| Contrast (Indirect 1 – Indirect 2) | −0.11 | 0.06 | −1.84 | .066 | −0.22, 0.00 |
| Total indirects (Indirect 1 + Indirect 2) | 0.20 | 0.05 | 4.32 | < .001 | 0.12, 0.31 |
| Total effects | 0.45 | 0.07 | 6.86 | < .001 | 0.33, 0.59 |

**Supplementary Table S6c.** Model coefficients assessing inward anger and impulsivity as parallel mediators from childhood adversity to self-aggression. Model is with age and gender as covariates.

| **Outcome / Path** | **β** | **SE (boot)** | **Z (boot)** | **p (boot)** | **95% CI (boot lower, upper)** | **R²** |
| --- | --- | --- | --- | --- | --- | --- |
| STAXI-2-axi: intercept (~1) | 3.12 | 0.39 | 7.95 | < .001 | 2.37, 3.92 | 0.05 |
| STAXI-2-axi ~ CTQ-tot (a1) | 0.21 | 0.08 | 2.84 | .004 | 0.05, 0.35 |  |
| STAXI-2-axi ~ Age | −0.02 | 0.10 | −0.19 | .852 | −0.19, 0.17 |  |
| STAXI-2-axi ~ Gender | 0.03 | 0.09 | 0.31 | .762 | −0.19, 0.21 |  |
| UPPS-urg: intercept (~1) | 4.71 | 0.42 | 11.12 | < .001 | 3.89, 5.59 | 0.23 |
| UPPS-urg ~ CTQ-tot (a2) | 0.25 | 0.08 | 3.16 | .002 | 0.11, 0.43 |  |
| UPPS-urg ~ Age | −0.34 | 0.08 | −4.32 | < .001 | −0.49, −0.17 |  |
| UPPS-urg ~ Gender | 0.03 | 0.09 | 0.33 | .743 | −0.15, 0.21 |  |
| KFAF-self: intercept (~1) | −0.14 | 0.53 | −0.26 | .793 | −1.20, 0.89 | 0.52 |
| KFAF-self~ STAXI-2-axi (b1) | 0.23 | 0.07 | 3.15 | .002 | 0.09, 0.37 |  |
| KFAF-self ~ UPPS-urg (b2) | 0.38 | 0.07 | 5.07 | < .001 | 0.23, 0.52 |  |
| KFAF-self~CTQ-tot (directc) | 0.18 | 0.07 | 2.59 | .010 | 0.05, 0.33 |  |
| KFAF-self ~ Age | −0.17 | 0.07 | −2.34 | .019 | −0.31, −0.03 |  |
| KFAF-self ~ Gender | 0.20 | 0.07 | 3.00 | .003 | 0.07, 0.33 |  |

| **Effect** | **β** | **SE (boot)** | **Z (boot)** | **p (boot)** | **95% CI (boot lower, upper)** |
| --- | --- | --- | --- | --- | --- |
| Direct (c) | 0.18 | 0.07 | 2.59 | .010 | 0.05, 0.33 |
| Indirect 1 (a1*b1) | 0.05 | 0.02 | 2.00 | .046 | 0.01, 0.10 |
| Indirect 2 (a2*b2) | 0.09 | 0.04 | 2.54 | .011 | 0.03, 0.18 |
| Contrast (Indirect 1 – Indirect 2) | −0.05 | 0.05 | −0.94 | .347 | −0.15, 0.04 |
| Total indirects (Indirect 1 + Indirect 2) | 0.14 | 0.04 | 3.46 | .001 | 0.08, 0.24 |
| Total effects | 0.32 | 0.07 | 4.72 | < .001 | 0.20, 0.47 |

**Supplementary Table S6d.** Model coefficients assessing inward anger and impulsivity as parallel mediators from childhood adversity to self-aggression. Model is with age, gender and cognitive depression severity as covariates.

| **Outcome / Path** | **β** | **SE (boot)** | **Z (boot)** | **p (boot)** | **95% CI (boot lower, upper)** | **R²** |
| --- | --- | --- | --- | --- | --- | --- |
| STAXI-2-axi: intercept (~1) | 2.80 | 0.43 | 6.52 | < .001 | 1.98, 3.67 | 0.09 |
| STAXI-2-axi ~ CTQ-tot (a1) | 0.14 | 0.09 | 1.57 | .117 | −0.05, 0.30 |  |
| STAXI-2-axi ~ Age | 0.06 | 0.11 | 0.56 | .572 | −0.13, 0.30 |  |
| STAXI-2-axi ~ Gender | −0.02 | 0.09 | −0.19 | .852 | −0.19, 0.16 |  |
| STAXI-2-axi ~ BDI-II-cog | 0.24 | 0.10 | 2.35 | .019 | 0.04, 0.44 |  |
| UPPS-urg: intercept (~1) | 4.28 | 0.43 | 9.94 | < .001 | 3.45, 5.16 | 0.30 |
| UPPS-urg ~ CTQ-tot (a2) | 0.16 | 0.08 | 1.87 | .062 | −0.01, 0.33 |  |
| UPPS-urg ~ Age | −0.24 | 0.08 | −2.83 | .005 | −0.40, −0.06 |  |
| UPPS-urg ~ Gender | −0.03 | 0.09 | −0.30 | .764 | −0.20, 0.15 |  |
| UPPS-urg ~ BDI-II-cog | 0.32 | 0.10 | 3.30 | .001 | 0.13, 0.51 |  |
| KFAF-self: intercept (~1) | −0.05 | 0.39 | −0.13 | .897 | −0.82, 0.72 | 0.72 |
| KFAF-self~ STAXI-2-axi (b1) | 0.16 | 0.06 | 2.81 | .005 | 0.05, 0.27 |  |
| KFAF-self ~ UPPS-urg (b2) | 0.23 | 0.06 | 4.00 | < .001 | 0.12, 0.35 |  |
| KFAF-self~CTQ-tot (directc) | 0.06 | 0.05 | 1.12 | .265 | −0.05, 0.16 |  |
| KFAF-self ~ Age | −0.03 | 0.06 | −0.52 | .606 | −0.15, 0.09 |  |
| KFAF-self ~ Gender | 0.11 | 0.05 | 2.02 | .043 | 0.01, 0.21 |  |
| KFAF-self ~ BDI-II-cog | 0.56 | 0.06 | 10.20 | < .001 | 0.45, 0.67 |  |

| **Effect** | **β** | **SE (boot)** | **Z (boot)** | **p (boot)** | **95% CI (boot lower, upper)** |
| --- | --- | --- | --- | --- | --- |
| Direct (c) | 0.06 | 0.05 | 1.12 | .265 | −0.05, 0.16 |
| Indirect 1 (a1*b1) | 0.02 | 0.02 | 1.35 | .176 | −0.01, 0.06 |
| Indirect 2 (a2*b2) | 0.04 | 0.02 | 1.57 | .118 | −0.00, 0.09 |
| Contrast (Indirect 1 – Indirect 2) | −0.01 | 0.03 | −0.43 | .665 | −0.08, 0.04 |
| Total indirects (Indirect 1 + Indirect 2) | 0.06 | 0.03 | 2.11 | .035 | 0.01, 0.12 |
| Total effects | 0.12 | 0.06 | 2.00 | .046 | −0.01, 0.24 |

***Note*** Table shows standardized estimates with bootstrap standard errors and bootstrap 95% confidence intervals based on 1,000 bootstrap samples. Z and p reported in the table are computed from the bootstrap SEs. R² values are reported for endogenous variables on their intercept rows. * p< .05; ** p< .01; *** p< .001. Gender is coded with female as reference.

Abbreviations: BDI-II-cog= cognitive subscore of the Beck Depression Inventory- Revision; CTQ-tot= total score of the Childhood Trauma Questionnaire; K-FAF= Kurzfragebogen zur Erfassung von Aggressivitätsfaktoren (Short Questionnaire for Assessing Factors of Aggression); STAXI-2= State-Trait Aggression Inventory second edition; UPPS= Urgency-Premeditation-Perseverance-Sensation Seeking questionnaire.

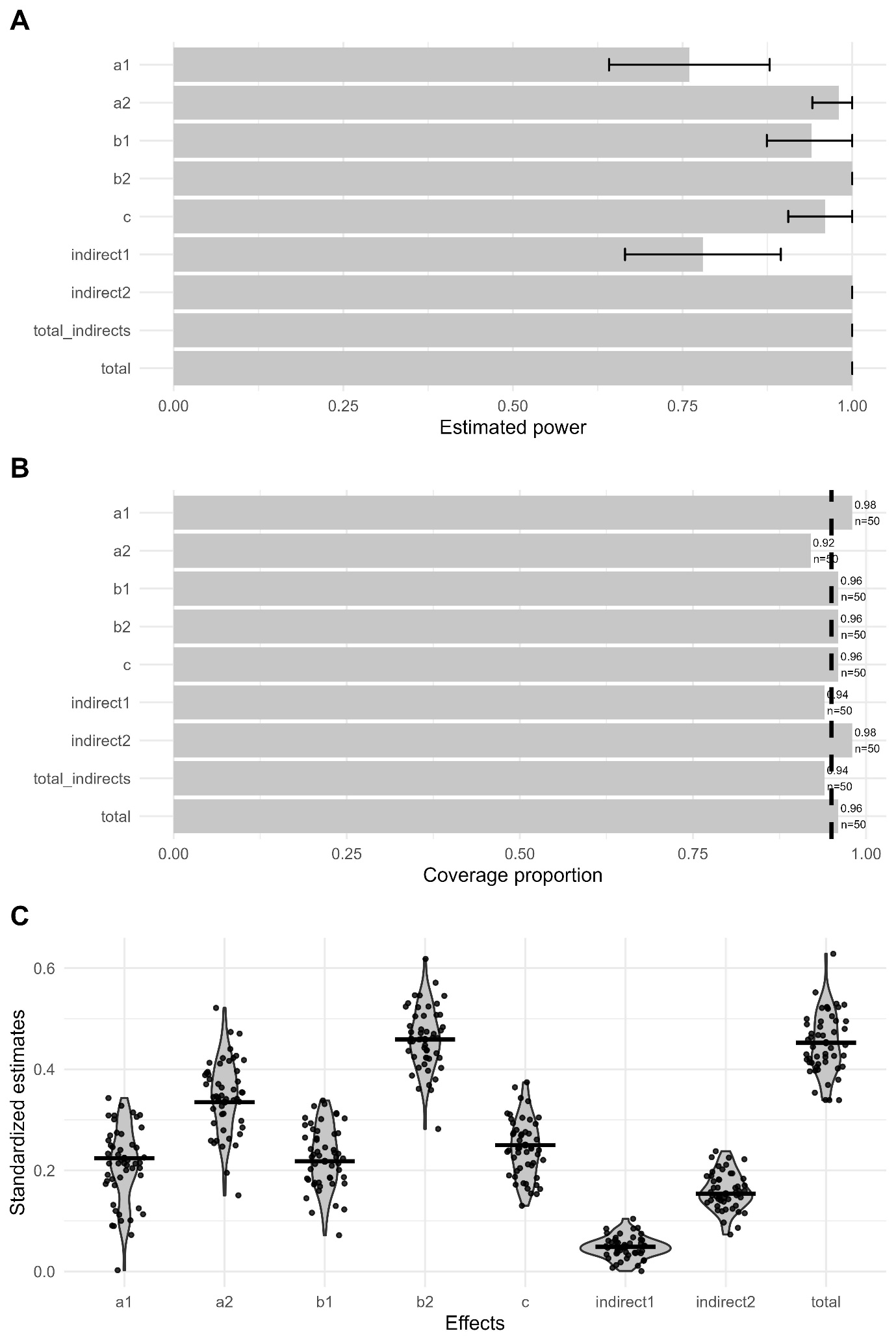

**Supplementary Figure 1** Figure accompanying power simulation for the parallel mediation model. A) Empirical power of effects with 95% MC CI. Bar plots show the proportion of successful simulated replications in which the bootstrap CI excluded zero (i.e., were significant) for indicated effects. The error bars are the 95% MC CI around those estimates. B) Coverage estimates. Bar plots display the proportion of simulated replications whose bootstrap CI contained the observed (herein reported) effect value; the dashed horizontal line marks the nominal 0.95 coverage. C) Distribution of standardized parameter estimates across simulations. Individual estimates from the simulations are displayed as jittered points while the horizontal black lines are the observed effect values.

***Note*** the graphs are for the n of simulations 250, with 1000 bootstrap resamples per fitted model. The details (proportion of significant replications, proportion of successful replications, power and 95% MC CI) for each effect are as follow: a1, ; a2, ; b1, ; b2, ; c, ; indirect1, ; indirect2, ; total indirects, ; total, .

Abbreviations: CI= confidence intervals; MC= Monte Carlo.
